# Ethnic differences in the manifestation of early-onset type 2 diabetes

**DOI:** 10.1101/2024.02.26.24303398

**Authors:** Davis Kibirige, Anita V. Hill, Jean Claude Katte, Isaac Sekitoleko, William Lumu, Julieanne Knupp, Steven Squires, Andrew T Hattersley, Liam Smeeth, Angus G. Jones, Moffat J. Nyirenda

**Author notes:** **Corresponding author** Davis Kibirige,. Joint senior authors (Angus G. Jones, Moffat J. Nyirenda).

## Abstract

**Aims/Hypothesis:** We undertook phenotypic characterization of early-onset and late-onset type 2 diabetes (T2D) in adult Black African and White European populations with recently diagnosed T2D to explore ethnic differences in the manifestation of early-onset T2D.

**Methods:** Using the Uganda Diabetes Phenotype study cohort of 500 adult Ugandans and the UK StartRight study cohort of 714 White Europeans with recently diagnosed islet autoantibody negative type 2 diabetes, we compared the phenotypic characteristics of participants with early-onset T2D (diagnosed at <40 years) and late-onset T2D (diagnosed at ≥40 years).

**Results:** One hundred and thirty four adult Ugandans and 113 White Europeans had early-onset T2D. Compared with late-onset T2D, early-onset T2D in White Europeans was significantly associated with a female predominance (52.2% vs. 39.1%, p=0.01), increased body mass index (mean [95% CI]-36.7 [35.2-38.1] kg/m^2^ vs. 33.0 [32.4-33.6] kg/m^2^, p<0.001), waist circumference (112.4 [109.1-115.6] cm vs. 108.8 [107.6-110.1] cm, p=0.06), and a higher frequency of obesity (82.3% vs. 63.4%, p<0.001). No difference was seen with the post-meal C-peptide levels as a marker of beta-cell function (mean [95% CI]-2130.94 [1905.12-2356.76] pmol/L vs. 2039.72 [1956.52-2122.92], p=0.62).

Conversely, early-onset T2D in Ugandans was associated with less adiposity (mean [95% CI] waist circumference-93.1 [89.9-96.3] cm vs. 97.4 [95.9-98.8] cm, p=0.006) and a greater degree of beta-cell dysfunction (120-minute post-glucose load C-peptide mean (95% CI) level-896.08 [780.91-1011.24] pmol/L vs 1310.10 [1179.24-1440.95] pmol/L, p<0.001), without female predominance (53.0% vs. 57.9%, p=0.32) and differences in the body mass index (mean [95% CI]-27.3 [26.2-28.4] kg/m^2^ vs. 27.9 [27.3-28.5] kg/m^2^, p=0.29).

**Conclusions/Interpretation:** These differences in the manifestation of early-onset T2D underscore the need for ethnic-specific therapeutic and preventive approaches for the condition.

**RESEARCH IN CONTEXT:** *What is already known about this subject?:*

- Globally, the burden of early-onset type 2 diabetes (T2D) is rapidly increasing.
- Evidence from studies conducted in Asians and White populations of European ancestry has shown that early-onset T2D is characterized by a female preponderance, increased obesity, rapid decline in the beta-cell function, and a high prevalence of diabetes-related complications.

*What is the key question?:*

- Does early-onset type 2 diabetes manifest differently in Black Africans and White populations of European ancestry with recently diagnosed type 2 diabetes?

*What are the new findings?:*

- Striking differences in the manifestation of early-onset type 2 diabetes were observed in adult Ugandans and White Europeans with recently diagnosed type 2 diabetes.
- Early-onset type 2 diabetes in adult White populations of European ancestry was characterized by female predominance and increased adiposity. In contrast, less adiposity, lower pancreatic beta-cell function, and absence of female predominance were observed in adult Ugandans with early-onset type 2 diabetes.

*How might this impact on clinical practice in the foreseeable future?:*

- Because of these phenotypic differences, the preventive and therapeutic strategies for early-onset type 2 diabetes should be individualized to ethnicity and population.

## INTRODUCTION

Globally, the burden of early-onset type 2 diabetes (T2D), defined as T2D diagnosed at <40 years, is rapidly increasing [1, 2]. Most of this evidence has been derived from Asian and White populations of European ancestry [3-8]. Data from these populations, particularly in high-income countries have shown that, compared with late-onset T2D, early-onset T2D is characterized by a female preponderance, increased obesity, rapid decline in the beta-cell function (reflecting an aggressive phenotype), and a high prevalence of diabetes-related complications [7, 9-13]. The condition is thought to develop due to a close interplay between genetics and environmental factors, with adiposity and genetic susceptibility as key features driving the early development of this condition [9, 10].

In contrast, evidence on the clinical and genetic profile of early-onset type 2 diabetes in adult Black African populations is limited [14, 15]. We also lack rigorous studies comparing the phenotypic characteristics of early-onset T2D (where islet-cell autoimmunity has been excluded) in Black adult Africans and White populations of European ancestry. To address this gap, we compared the demographic, clinical, anthropometric, and metabolic characteristics of adult Ugandans and White Europeans with early-onset T2D (diagnosed at <40 years) and late-onset T2D (diagnosed at ≥40 years) recruited in the Uganda Diabetes Phenotype (UDIP) and the United Kingdom StartRight studies, respectively, with an overarching aim of exploring whether differences in the manifestation of early-onset T2D exist between these two distinct populations.

## METHODS

The participants in this study were recruited in the UDIP and StartRight studies conducted in Uganda and the United Kingdom (UK), respectively. The UDIP study aimed to undertake rigorous phenotypic characterization of 568 adult Ugandans (aged ≥18 years) with recently diagnosed diabetes (diabetes diagnosed in the preceding three months). The study participants were recruited from February 2019 to October 2020 from the adult diabetes outpatient clinics of seven public and faith-based private not-for-profit secondary hospitals in Central and Southwestern Uganda. All pregnant women with recently diagnosed diabetes were excluded from the study.

The StartRight study was a prospective multi-center study conducted across 55 sites in the UK that aimed to provide robust clinical evidence on accurate classification of diabetes at the time of diagnosis and early identification of patients who will rapidly require insulin treatment (https://www.clinicaltrials.gov/study/NCT02287506). The study recruited 1,802 participants aged ≥ 18 years with a diagnosis of diabetes made within the previous 12 months. Participants with gestational and secondary diabetes were excluded from the study. The StartRight study was enriched for late-onset type 1 diabetes by aiming for equal recruitment of those receiving and not receiving insulin therapy in those with diabetes onset after 50 years. For our analysis, we considered only individuals of White ancestry with a clinical diagnosis of type 2 diabetes and confirmed negative islet autoantibody status.

### Assessment of the phenotypic characteristics of interest

Regarding the UDIP study, information on the demographic (age, sex, and residence) and clinical (family history of diabetes and history of co-existing hypertension) characteristics of interest was collected from each study participant. This was followed by resting blood pressure and anthropometric measurement and the systolic and diastolic blood pressure, weight, height, waist circumference (WC), hip circumference (HC), BMI, and waist: hip circumference ratio (WHR) were recorded.

A fasting venous blood sample was then drawn for the measurement of glycated hemoglobin (HbA1c), lipid profile, C-peptide, and three islet autoantibodies (glutamic acid decarboxylase-65 or GADA, tyrosine phosphatase or IA-2A, and zinc transporter 8 or ZnT8-A). All participants were subjected to a 75-gram oral glucose tolerance test (OGTT) to measure the 120-minute C-peptide concentrations. All the above tests were carried out at the Medical Research Council/Uganda Virus Research Institute and London School of Hygiene and Tropical Medicine Uganda Research Unit, Entebbe Uganda.

For the StartRight study, demographic and clinical information was collected at the recruitment visit with an assessment of height, weight, waist, and hip circumference measurements to calculate the BMI and WHR. This was followed by the collection of a nonfasted (within 1-5 hours post-meal) blood sample for DNA extraction, and measurement of C-peptide, glucose, HbA1c, and islet autoantibodies (GADA, IA-2A, and ZnT8A) concentrations. These tests were performed at the academic Blood Sciences Department at the Royal Devon and Exeter Hospital, Exeter UK.

### Assessment of type 1 and type 2 diabetes Genetic Risk Score

In addition to screening for the three islet autoantibodies, we calculated type 1 diabetes (T1D) and T2D genetic risk scores (GRS). The T1D GRS was calculated based on the 67 Single Nucleotide Polymorphisms (SNPs) reported to be associated with T1D, as described by Sharp et al [16]. The T2D GRS was calculated using T2D-associated SNPs from a European population [17] which was shown to effectively predict T2D across racial/ethnic groups [18].

### Definition of the study outcomes

Early-onset and late-onset T2D were defined as T2D diagnosed at <40 years and ≥40 years, respectively, with confirmed islet autoantibody negative status in both populations. Ugandan participants were considered to be islet autoantibody negative if the GADA, IA-2A, and ZnT8-A concentrations were ≤34 U/ml, ≤58 U/ml, and ≤67.7 U/ml, respectively, based on the 97.5^th^ centile of 600 adult rural Ugandans without diabetes.

Islet autoantibody negative status was considered in the White European participants if the GADA and IA-2A concentrations were <11 U/ml and <7.5 U/ml, respectively. Cut-offs of <65 U/ml and <10 U/ml were used for the ZnT8-A concentration in participants aged <30 years and ≥30 years, respectively [19]. These cut-offs represented the ≥97.5^th^ percentile of 1,559 British participants without diabetes.

Obesity was defined based on the traditional World Health Organisation cut-off of ≥30 kg/m^2^.

### Statistical analysis

Percentages and medians with their corresponding interquartile range (IQR) were used to describe the categorical and continuous variables, respectively. The demographic, clinical, anthropometric, and metabolic characteristics of the participants with early-onset and late-onset T2D were analyzed using Fisher’s exact test for categorical data and the two-sample t-tests for continuous data. The categorical and continuous variables were expressed as proportions and mean with 95% confidence intervals (CI), respectively. All analyses were performed using STATA statistical software version 15 (StataCorp, USA).

### Ethical approval

This UDIP study was approved by the Research Ethics Committee of Uganda Virus Research Centre, Entebbe Uganda (GC/127/18/05/650) and the Uganda National Council of Science and Technology (HS 2431), with administrative approval from all participating study sites. The StartRight study was approved by the South West– Cornwall and Plymouth NHS Research Ethics Committee (16/SW/0130). All enrolled study participants provided written informed consent to participate in the study.

## RESULTS

### Characteristics of the White European and Ugandan participants with islet autoantibody negative type 2 diabetes

The overall characteristics of the White European and Ugandan participants with islet autoantibody negative type 2 diabetes are summarized in Table 1.

**Table 1.** Overall characteristics of the adult White Europeans and Black Ugandans with recently diagnosed type 2 diabetes.

| Characteristics | Adult White European participants with recently diagnosed T2D (n= 714) | Adult Black Ugandan participants with recently diagnosed T2D (n= 500) |
| --- | --- | --- |
| Age at diagnosis (years) | 51.2 (50.2-52.2) | 48.4 (47.2-49.5) |
| Female, n (%) | 294 (41.2%) | 283 (56.6%) |
| Parental history of T2D, n (%) | 318 (44.5%) | 191 (38.2%) |
| Co-existing hypertension, n (%) | 268 (37.5%) | 171 (35.0%) |
| Systolic blood pressure, mmHg | 134.4 (133.1-135.7) | 126.8 (125.2-128.0) |
| Diastolic blood pressure, mmHg | 81.0 (80.2-81.8) | 84.1 (83.1-85.0) |
| Body mass index, kg/m<sup>2</sup> | 33.6 (33.0-34.2) | 27.7 (27.2-28.2) |
| Body mass index ≥30 kg/m<sup>2</sup>, n (%) | 474 (66.4%) | 171 (34.2%) |
| Waist circumference, cm | 109.4 (108.2-110.6) | 96.3 (94.9-97.6) |
| Hip circumference, cm | 113.6 (112.5-114.6) | 104.2 (102.3-106.1) |
| Waist: hip ratio | 0.96 (0.96-0.97) | 0.92 (0.91-0.93) |
| TC, mmol/l | 5.20 (5.05-5.35) | 4.17 (4.05-4.29) |
| HDLC, mmol/l | 1.18 (1.13-1.24) | 0.95 (0.74-1.20) |
| LDLC, mmol/l | 2.95(2.83-3.06) | 2.71 (2.61-2.81) |
| TGL, mmol/l | 3.40 (2.88-3.93) | 1.59 (1.50-1.67) |
| HbA1c, mmol/mol | 73.2 (71.2-75.2) | 88.2 (85.0-91.3) |
| HbA1c, % | 8.8 (8.7-9) | 10.2 (9.9-10.5) |
| Random/120-minute post-OGTT C-peptide, pmol/L * | 2054.16 (1975.73-2132.58) | 1197.79 (1096.34-1299.25) |

Continuous data is expressed as mean (95% CI), \*Random or nonfasted, and 120- minute post-OGTT load C-peptide concentrations used for the White European and Ugandan participants, respectively.

Compared with the White European participants, adult Ugandans with islet autoantibody negative type 2 diabetes were younger (mean [95% CI]-48.4 [47.2-49.5] years vs. 51.2 [50.2-52.2] years) with lower markers of adiposity (mean [95% CI] BMI of 27.7 [27.2-28.2] kg/m^2^ vs. 33.6 [33.0-34.2] kg/m^2^, and BMI ≥30 kg/m^2^ - 34.2% vs. 66.4%). Additionally, Ugandan participants had a higher HbA1c (mean [95% CI] HbA1c – 88.2 [85.0-91.3] mmol/mol vs. 73.2 [71.2-75.2] mmol/mol), and lower pancreatic beta-cell function (mean [95% CI] post-meal or 120-minute post-glucose load C-peptide-1197.79 [1096.34-1299.25] pmol/L vs. 2054.16 [1975.73-2132.58] pmol/L) at the time of recruitment.

### In White Europeans, early onset T2D is strongly associated with a female predominance and markers of obesity

The characteristics of the adult White European and Ugandan participants with early-onset and late-onset T2D are shown in Table 2 and Figure 1.

**Table 2.** Characteristics of adult White European and Ugandan participants with early-onset and late-onset type 2 diabetes.

| Characteristics | Adult White European population with recently diagnosed type 2 diabetes (n= 714) |  |  | Adult Black Ugandan population with recently diagnosed type 2 diabetes (n= 500) |  |  |
| --- | --- | --- | --- | --- | --- | --- |
|  | <40 years<br>(n= 113, 15.8%) | ≥40 years<br>(n= 601, 84.2%) | P-value | <40 years<br>(n= 134, 26.8%) | ≥40 years<br>(n= 366, 73.2%) | P-value |
| Age at diagnosis (years) | 32.7 (31.7-33.7) | 54.7 (53.8-55.5) |  | 32.9 (32.0-33.8) | 54.0 (53.0-55.0) |  |
| Female, n (%) | 59 (52.2%) | 235 (39.1%) | 0.01 | 71 (53.0%) | 212 (57.9%) | 0.32 |
| Parental history of T2D, n (%) | 56 (49.6%) | 262 (43.6%) | 0.26 | 54 (40.3%) | 137 (37.4%) | 0.68 |
| Co-existing HT, n (%) | 12 (10.6%) | 256 (42.6%) | <0.001 | 16 (11.9%) | 155 (42.4%) | <0.001 |
| Systolic BP, mmHg | 127.8 (125.3-130.3) | 135.6 (134.2-137.0) | <0.001 | 120 (117-122) | 129 (127-131) | <0.001 |
| Diastolic BP, mmHg | 80.2 (78.4-82.0) | 81.1 (80.3-82.0) | 0.27 | 81 (79-83) | 85 (84-86) | <0.001 |
| BMI, kg/m<sup>2</sup> | 36.7 (35.2-38.1) | 33.0 (32.4-33.6) | <0.001 | 27.3 (26.2-28.4) | 27.9 (27.3-28.5) | 0.29 |
| BMI ≥30 kg/m<sup>2</sup>, n (%) | 93 (82.3%) | 381 (63.4%) | <0.001 | 43 (32.6%) | 128 (35.2%) | 0.48 |
| Waist circumference, cm | 112.4 (109.1-115.6) | 108.8 (107.6-110.1) | 0.06 | 93.1 (89.9-96.3) | 97.4 (95.9-98.8) | 0.006 |
| Hip circumference, cm | 119.4 (116.5-122.3) | 112.5 (111.4-113.6) | <0.001 | 104.2 (98.4-110.0) | 104.2 (102.7-105.7) | 0.99 |
| Waist: hip ratio | 0.94 (0.93-0.96) | 0.97 (0.96-0.98) | 0.003 | 0.89 (0.88-0.91) | 0.93 (0.92-0.94) | <0.001 |
| Total cholesterol, mmol/l | 5.65 (4.97-6.32) | 5.12 (5.00-5.25) | 0.17 | 4.03 (3.83-4.20) | 4.22 (4.08-4.36) | 0.17 |
| HDLc, mmol/l | 1.04 (0.96-1.13) | 1.21 (1.15-1.27) | 0.002 | 0.94 (0.70-1.15) | 0.96 (0.76-1.23) | 0.52 |
| LDLC, mmol/l | 3.11 (2.84-3.39) | 2.92 (2.80-3.05) | 0.22 | 2.56 (2.38-2.73) | 2.76 (2.64-2.88) | 0.07 |
| Triglycerides, mmol/l | 3.87 (1.68-6.06) | 3.33 (2.83-3.83) | 0.29 | 1.54 (1.39-1.70) | 1.60 (1.50-1.71) | 0.55 |
| HbA1c, mmol/mol | 77.6 (72.5-82.8) | 72.4 (70.2-74.5) | 0.049 | 96.3 (89.7-102.9) | 85.3 (81.7-88.8) | 0.004 |
| HbA1c, % | 9.3 (8.8-9.7) | 8.8 (8.6-9.0) | 0.049 | 10.9 (10.3-11.5) | 9.9 (9.6-10.3) | 0.004 |
| Random/120-minute post-OGTT C-peptide, pmol/L* | 2130.94 (1905.12-2356.76) | 2039.72 (1956.52-2122.92) | 0.62 | 896.08 (780.91-1011.24) | 1310.10 (1179.24-1440.95) | <0.001 |
| T1D GRS | 10.42 (9.93-10.92) | 10.07 (9.88-10.26) | 0.14 | 8.43 (8.03 – 8.81) | 8.12 (7.89 - 8.36) | 0.20 |
| T2D GRS | 11.25 (11.19-11.31) | 11.20 (11.17-11.22) | 0.08 | 11.28 (11.21-11.34) | 11.22 (11.18-11.25) | 0.09 |

Continuous data is expressed as mean (95% CI), \*Random or nonfasted and 120-minute post-OGTT load C-peptide concentrations used for the White European and Ugandan participants, respectively.

**Figure 1.**
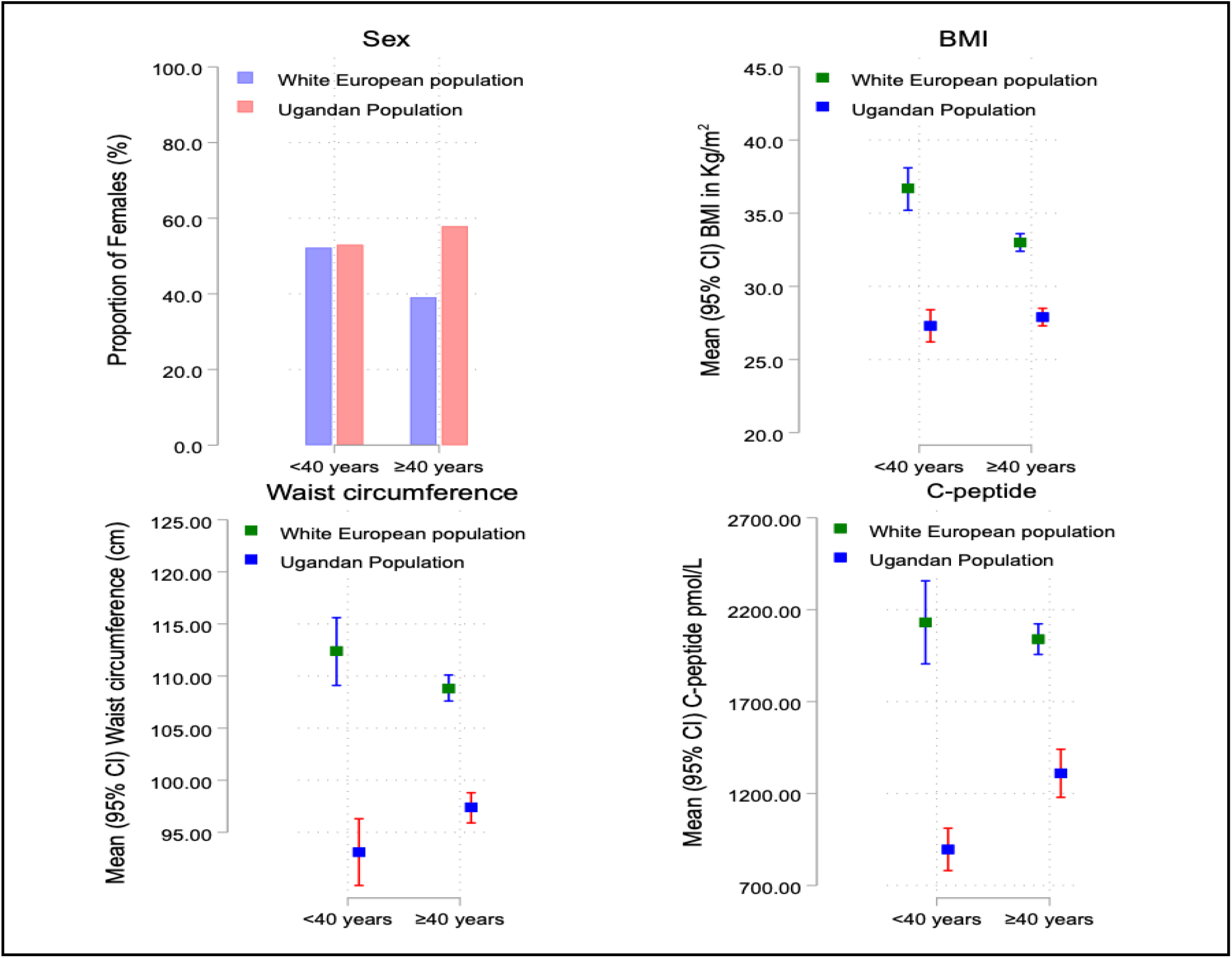
Comparison of the female sex distribution, body mass index, waist circumference, and post-glucose or meal C-peptide levels between the White European and Ugandan participants with early-onset and late-onset type 2 diabetes. BMI-Body mass index

Of the 500 Ugandan and 714 White European participants enrolled in the UDIP and UK StartRight studies with autoantibody negative T2D, 134 (26.8%) and 113 (15.8%) had early-onset T2D, respectively.

Compared with late-onset T2D, early-onset T2D in the White European participants was significantly associated with a female predominance (52.2% vs. 39.1%, p=0.01), increased BMI (mean [95% CI]-36.7 [35.2-38.1] kg/m^2^ vs. 33.0 [32.4-33.6] kg/m^2^, p<0.001), WC (112.4 [109.1-115.6] cm vs. 108.8 [107.6-110.1] cm, p=0.06), and a higher frequency of obesity (82.3% vs. 63.4%, p<0.001).

Conversely, compared with those with late-onset T2D, Ugandan participants with early-onset T2D had lower markers of adiposity (mean [95% CI] WC-93.1 [89.9-96.3] cm vs. 97.4 [95.9-98.8], p=0.006). No differences were noted in the proportion of females (53.0% vs. 57.9%, p=0.32), frequency of obesity (32.6% vs. 35.2%, p=0.48), and BMI levels (mean [95% CI]- 27.3 [26.2-28.4] kg/m^2^ vs. 27.9 [27.3-28.5] kg/m^2^) in Ugandan participants with early-onset and late-onset T2D.

### Early onset T2D in Ugandans is associated with marked hyperglycemia and pancreatic beta cell failure

Ugandan participants with early-onset T2D had a greater degree of hyperglycemia (mean [95% CI] HbA1c-96.3 [89.7-102.9] mmol/mol vs 85.3 [81.7-88.8] mmol/mol, p=0.004) and pancreatic beta-cell dysfunction (mean [95% CI] 120-minute post-glucose load C-peptide level-896.08 [780.91-1011.24] pmol/L vs 1310.10 [1179.24-1440.95] pmol/L, p<0.001) at the time of recruitment. In contrast, the post-meal C-peptide levels did not differ in the White European participants with early-onset and late-onset T2D (mean [95% CI]- 2130.94 [1905.12-2356.76] pmol/L vs. 2039.72 [1956.52-2122.92] pmol/L, p=0.62).

### Genetic risk score of the adult Ugandan and White European participants with early-onset type 2 diabetes

Ugandan participants with early-onset T2D had a higher T2D GRS when compared with those with late-onset T2D (11.28 [11.21-11.34] vs. 11.22 [11.18-11.25], p=0.09). Similarly, compared with those with late-onset T2D, White European participants with early-onset T2D also had a higher T2D GRS (11.25 [11.19-11.31] vs. 11.20 [11.17-11.22], p=0.08), although the differences in both cohorts were not statistically significant.

## DISCUSSION

To our knowledge, this is the first study to rigorously investigate the manifestation of early-onset and late-onset T2D in an adult Black African population and to make comparisons with a White European population. We demonstrate striking differences in presentation between these two populations, where, in contrast to the White European cohort, early-onset T2D in adult Ugandans is likely to be seen more in patients without obesity (and other features of the metabolic syndrome), and is associated with marked pancreatic beta-cell dysfunction.

The association between obesity and early-onset T2D in White European populations has been well documented and suggests a common underlying mechanism, driven by insulin resistance as a primary defect and pancreatic beta-cell dysfunction occurring later [4, 20-22]. However, beta-cell failure appears to occur more rapidly in patients with early-onset T2D compared with those with late-onset T2D [9, 11]. In addition, early-onset T2D in White European participants was common among females, which has also been widely reported in other studies of White populations [23, 24].

In contrast, the pathways that lead to the early onset of T2D in Africa, where it occurs in the absence of excessive adiposity, are unclear, but most evidence suggests that pancreatic beta-cell dysfunction is the primary defect. This may result from genetic predisposition as well as environmental exposures, such as early-life (in-utero and/or early childhood) malnutrition and infections like tuberculosis, HIV, and malaria, which are prevalent in the region [25-28]. This notion forms the basis of the developmental origins of health and disease (DOHaD), which explains that such early-life environmental exposures may induce changes, including epigenetic, that alter gene expression, cellular growth, composition, and physiology, increasing the future risk of developing cardiometabolic conditions like T2D [29]. The high burden of infectious diseases, such as tuberculosis and HIV, may also act directly or indirectly in adulthood to increase the risk of developing early-onset T2D [25, 26].

The unique manifestation of early-onset T2D with a predominance of pancreatic beta-cell dysfunction, as seen in adult Ugandans, may also be due to genetic influences. Polymorphisms of genes that influence pancreatic beta-cell development, proliferation, neogenesis, apoptosis, and insulin secretion such as the transcription factor-7 like 2 genes (TCF7L2), Zinc Finger RANBP2-Type Containing 3 (*ZRANB3*), and the ATP-sensitive potassium channel Kir6.2 gene (KCNJ11), have been suggested as possible mechanisms for pancreatic beta-cell dysfunction in patients of African ancestry [30-32].

Strengths of the study include that this is the first study to undertake phenotypic characterization of early-onset and late-onset T2D (where islet-cell autoimmunity has been robustly screened for and excluded) in adult Black African and White European patients with recently diagnosed diabetes, to investigate if ethnic-related differences exist in the manifestation of early-onset T2D. Participants were phenotyped in detail, and GRS was available to investigate the potential contribution of antibody-negative T1D and classical T2D genetic susceptibility to early-onset antibody-negative T2D. Limitations of this study include differences in the designs of the two studies which may limit the interpretation of some findings. For example, the differences in the study design (including age-related recruitment in the StartRight study) suggest that these studies should not be used to directly compare the relative prevalence of early-onset T2D. In addition, the post-meal C-peptide measurement in the StartRight study may not be directly comparable to the 120-minute post-glucose load C-peptide measurement used in the UDIP study, and differences in duration at recruitment may impact direct comparison of glycemic control. Ugandan participants were recruited only from seven secondary hospitals (in contrast to the combined primary and secondary healthcare recruitment in the StartRight study).

In conclusion, our study findings demonstrate that the phenotypic profile of early-onset T2D in adult Ugandans and White Europeans with recently diagnosed diabetes greatly differs. While obesity plays a central role in the pathogenesis of early-onset T2D in White Europeans, its effect in adult Ugandans is insignificant. Pancreatic beta-cell dysfunction appears to explain the early onset of T2D in this population. An in-depth understanding of the phenotype of early-onset T2D in Black African and White European populations is important and has significant clinical and therapeutic implications. Because of these phenotypic differences, the therapeutic and preventive strategies for early-onset T2D should be tailored to ethnicity and population. Due to a lack of adequate clinical evidence, future research is needed to guide how to optimally manage and prevent early-onset T2D, especially in adult Ugandans.

## Data Availability

All data produced in the present study are available upon reasonable request to the authors

## Acknowledgments

The authors are grateful to all the study participants who consented to join the study, the UDIP and StartRight research teams, the staff of the Clinical Diagnostics Laboratory Services at the Medical Research Council/Uganda Virus Research Institute and London School of Hygiene and Tropical Medicine Uganda Research Unit, Entebbe, Uganda, and the academic Blood Sciences Department, Royal Devon and Exeter Hospital, UK, for conducting the laboratory tests. The authors also thank the ADDRESS-2 study team (Imperial College, London, UK) for support with participant recruitment in the StartRight study.

## Data availability statement

The datasets generated during and/or analyzed in the current study are available from the corresponding author upon reasonable request.

## Funding and Assistance

This UDIP study was supported by the UK Medical Research Council (MRC) and the UK Department for International Development (DFID) under the MRC/DFID Concordat agreement (Project Reference: MC_UP_1204/16), and the National Institute for Health Care Research (NIHR) (17/63/131). The StartRight study was funded by the National Institute for Health and Care Research (NIHR) (CS-2015-15-018) and Diabetes UK (17/0005624). Genetic analysis for the StartRight study was funded by the European Foundation for the Study of Diabetes (2016 Rising Star Fellowship).

JCK is supported by the NIHR Exeter Biomedical Research Centre (NIHR Exeter BRC). ATH is supported by the NIHR Exeter Clinical Research Facility and an NIHR Senior Investigator award. AGJ was supported for this work by an NIHR Clinician Scientist award (CS-2015-15-018). MJN is an MRC Investigator.

The study sponsor/funder was not involved in the design of the study; the collection, analysis, and interpretation of data; writing the report; and did not impose any restrictions regarding the publication of the report.

## Authors’ relationships and activities

No potential conflicts relevant to this article were reported.

## Contribution statement

DK and JCK wrote the first draft of the manuscript, DK and AVH participated in the study design, data acquisition, analysis, and interpretation, JCK, IS and JK participated in the study design, data analysis, and interpretation, and reviewed all the versions of the manuscript, WL participated in the data collection process, and data interpretation, SS performed the genetic analysis and reviewed all the versions of the manuscript, AGJ, ATH, LS, and MJN supervised this work, received funding for the entire research project, reviewed, and edited all versions of the manuscript. All authors read and approved the final draft of the manuscript. DK is the guarantor of this work and, as such, had full access to all the data in the study and takes responsibility for the integrity of the data and the accuracy of the data analysis.

